# Physiotherapy Under Pressure: A Cross-Sectional Study on The Interplay Between Perfectionism, Moral Injury, and Burnout

**DOI:** 10.1101/2024.02.09.24302501

**Authors:** Daniel Biggs, Laura Blackburn, Cameron Black, Sivaramkumar Shanmugam

## Abstract

**Background:** Given the escalating challenges for UK-based physiotherapists in workload pressures, budget constraints, staff shortages and patient wait times, the profession (of 65,000 registered physiotherapists) requires immediate care and review of therapist health. This pioneering study aims to examine perfectionism, moral injury, and burnout among UK-based physiotherapists across the NHS, private practice, sports, and academia.

**Method:** This cross-sectional study utilised an online survey and implementation of Structure Equation Modelling (SEM) to assess the interplay of Perfectionism (Multidimension Perfectionism Scale-Short Form), Moral Injury (Moral Injury Symptoms Scale-Healthcare Professionals) and Burnout (Shirom-Melamed Burnout Questionnaire). Our sample size calculation shows representation of the UK physiotherapy profession via utilisation of 95% confidence interval with a 5% margin of error.

**Findings:** Our analysis conducted on (n=402) UK-based physiotherapists reveals significant burnout levels – 96% are categorised as moderate to high. Furthermore, associations and SEM of Perfectionism and Moral Injury collectively account for a substantial proportion of burnout variability (r^2^.62), highlighting their sequential impact on burnout manifestation.

**Interpretation:** With such high levels of burnout, urgent intervention is paramount. Elevated burnout presents challenges for the physiotherapy profession as staff retention, accurate and effective patient care, and overall health are severely impacted due to burnout. Recognising and addressing perfectionism and moral injury becomes pivotal to mitigate its impact on individual and collective health.

**Funding:** No funding was acquired for this research project.

## Introduction

### Perfectionism

Bayeux Tapestry encapsulates the essence of perfectionism through its intricate embroidery process, where each thread is meticulously woven to depict mediaeval scenes with precision. Embroiderers become immersed in a meticulous battle, fixated on capturing every intricate detail to ensure the tapestry emerges as a flawless masterpiece. Parallels between Bayeux Tapestry and perfectionism extends beyond the meticulous process to encompass the high standards of the creators. In both cases, there is commitment to achieving absolute flawlessness, be it the careful stitching of threads or the pursuits of perfection in one’s work. Bayeux Tapestry thus serves as a tangible representation of the relentless pursuit of flawlessness, mirroring the psychology of perfectionism. Commonly acknowledged as an intricate multifaceted personality trait, perfectionism can be characterised by exceedingly high levels of self-critical judgement and personal standards, encompassing a synthesis of rigorous self-evaluative standards.^1^

Frost et al^2^ outlined six dimensions of perfectionism. These encompassed Personal Standards, characterised by high internal expectations; Concerns Over Mistakes, reflecting apprehensions about errors; Parental Criticism, entailing critical evaluations/input from parents; Parental Expectations, involving demands of perceived flawlessness form parents; Doubts About Actions, encapsulating subjective uncertainty about performance; and Organisation, emphasising preparedness and methodicalness. The framework explores elevated self-expectations, maladaptive cognitive patterns related to mistakes, inward perfectionism, perceived parental pressure for flawlessness, subjective uncertainty about performance, and the acknowledgement of the significance of preparedness and methodical approaches. An alternative perspective on perfectionism emerged from Hewitt and Flett’s model^3^, which also posits perfectionism as multidimensional, but with three instead of six dimensions. These dimensions are Self Oriented Perfectionism (SOP), characterised by an inherent drive for flawlessness leading to self-criticism and high personal standards; Socially Prescribed Perfectionism (SPP), arising from external expectations form significant others like parents, guardians, coaches or teachers, resulting in excessive self-criticism when unmet; and Other Oriented Perfectionism (OOP), revolving around expecting perfection from others and manifesting in hyper-critical behaviour. The present study adopts the perspective of perfectionistic standards being viewed by the individual from the self and therefore renders OOP redundant to investigate with moral injury and burnout. Despite disparity in dimensions, common themes resonate between the two models, such as the imposition of high standards in both internal and external contexts and the presence of maladaptive cognitions during the pursuit and outcomes of goals. Literature favours Hewitt and Flett’s model, drawn by its validation through factor analysis, supporting the existence of adaptive and maladaptive facets of perfectionism.^4^

A recent review focused on SPP, describing it as an epidemic.^5^ Flett et al^5^ showed the destructive nature of SPP through links with poor mental well-being, physical health, interpersonal adjustment, suicidal cognitions and tendencies, and burnout. The review stated, “SPP is a significant public health concern that urgently requires sustained prevention and intervention efforts”.^5(p1)^ As SPP is a pressure to meet external standards prescribed by the social environment, it may be problematic in certain settings and job roles, particularly when multiple factors contribute to decision-making. In the realm of healthcare, UK-based physiotherapists operate as independent practitioners across various sectors, such as public, private or sports settings. It is noteworthy that a significant majority, approximately 70% of UK-based physiotherapists work in the NHS.^6^ While these practitioners enjoy autonomy in their roles, it is important to recognise their independence is circumscribed by NHS procedures. Consequently, physiotherapists must carefully consider numerous factors when devising treatment plans. This poses a challenge for the physiotherapist, who must navigate the delicate balance of meeting the needs of patients whilst adhering to organisational demands. In essence, the practitioner is tasked with harmonising the diverse demands and standards of both the patient and the organisation. However, this intricate balancing act may introduce elements of unpredictability for the physiotherapist, with discrepancies arising among the therapist, patient, and the organisation, potentially compromising the practitioner’s own professional needs and the innate perfection in their personality.

### Moral Injury

Moral injury, rooted in the ethical branch of axiology and philosophy, revolves around an individual’s view on what is right and what is wrong.^7^ This form of injury often occurs when there is a perceived betrayal of moral integrity, from either someone of legitimate authority or oneself during a high-stakes situation.^8^ Moral injury renders feelings of shame, remorse, meaninglessness, and grief due to the violation of core moral beliefs.^9^ This deeply personal experience often shapes one’s identity and the recognition of a breach in these beliefs becomes evident through the resultant moral injury. A growing body of literature explores moral injury in healthcare professionals. The oath taken by healthcare practitioners, prioritising patient needs, serves as a guiding principle for their conduct and decisions.^10^ However, competing factors such as insurance considerations, hospital dynamics, electronic medical records, the healthcare system model, and personal financial security can take precedence over patient well-being. Over time, decisions deviating from the best interests of patients can be perceived as a moral injustice, leading to feelings of shame and guilt. These emotions, in turn, may contribute to a desire to leave the healthcare profession.^11^ This parallels the concept of SPP, where the pursuit of flawless decisions and standards may be hindered by external factors, making the attainment of perfection seemingly unachievable.

Čartolovni et al^12^ presents an alternative perspective to Dean and colleague’s findings^10^, highlighting healthcare professionals encounter moral injury due to the discord between their duty to provide care, their fundamental professional role, and the witnessing of traumatising events that impose an ethical burden on them. This significantly impacts the mental health of healthcare professionals. Instances of trauma include the inadequate allocation of resources, a pervasive sense of hopelessness and helplessness in overcrowded hospitals, and the emotional toll of witnessing innocent deaths, resulting from moral decision-making in triage prioritisation.^12^ This underscores that moral injury is not a uniform experience; rather, healthcare professionals face vulnerability arising from various aspects of their roles in the job.

Moral injury must be clearly distinguished from Post-Traumatic Stress Disorder (PTSD), this differentiation is crucial in the healthcare context, where the focus lies on understanding how moral injury arises, from the frustration experienced by healthcare professionals. Although these individuals are trained to provide care, they encounter barriers leading to an inability to deliver the expected care. This discrepancy can result in various adverse psychological outcomes, including burnout. Notably, research has demonstrated a correlation between burnout and moral injury.^13^ Recognising and appreciating these distinctions is important for healthcare practitioners and researchers, as it sheds light on the unique challenges posed by moral injury, and the resultant repercussions in healthcare settings.

### Burnout

A syndrome with far-reaching consequences, burnout affects not only the well-being and performance of individuals but also organisational outcomes, such as employee turnover, absenteeism, and diminished productivity.^14^ Three core dimensions of exhaustion, cynicism, and loss of commitment albeit with subtle changes are conceptualised by Maslach and Jackson.^15^ These authors show burnout as a tri-dimensional syndrome, encompassing Emotional and Psychophysical Exhaustion, linked to somatic-like symptoms, such as tiredness or headaches; Depersonalisation, involving the tendency to detach oneself from society and societal values; and Reduced Sense of Accomplishment, indicating an adverse appraisal of individual abilities or achievements. Since the inception of this model, various others have been developed and grounded in the three dimensions of Maslach and Jackson’s model.^15^

The Shirom-Melamed Burnout Measure (SMBM) for has been used in various studies.^16^ Validated through confirmatory factor analysis^16^, the SMBM is characterised by three dimensions: emotional exhaustion (EE), physical fatigue (PE) and cognitive weariness (CW). Whether researchers employ Maslach and Jackson’s model or the SMBM model, burnout onset is influenced by a range of factors at the individual (micro), occupational (meso), and organisational (macro) levels. These factors, encompassing high job demands, insufficient resources and support, a lack of autonomy and control, role ambiguity, and interpersonal conflicts are further influenced by the interaction of personality traits, coping styles, and prior experiences, shaping the emergence and progression of burnout.^17^

Donohoe et al^18^ pioneered the measurement of burnout in physiotherapists, revealing 40% experienced moderate-to-high levels of burnout. Subsequent studies consistently delved into the phenomenon of burnout in physiotherapists, reinforcing the vulnerability of physiotherapists to burnout.^19^ It is important to note these studies span various countries with different healthcare systems, and the impact of contributing factors may vary across contexts. Nevertheless, the collective evidence indicates burnout is a prevalent and significant issue among physiotherapists, reaching moderate-to-high levels across diverse settings.

Literature on burnout in physiotherapists spans different countries and continues to centre on prevalence. However, there is a notable absence of research on burnout in UK physiotherapists. Investigating this aspect is crucial for advancing our understanding and allowing for international and interdisciplinary comparisons. It also provides an opportunity to examine burnout within the context of the UK healthcare system, particularly post-pandemic.

### Perfectionism, Moral Injury and Burnout

Research highlights links between perfectionism, moral injury, and burnout.^13,20^ For example, Testoni et al^13^ found a relationship (r .20 p = <.001) between moral injury and burnout in clinicians. This finding is important as clinicians tend to dehumanise patients and colleagues during high levels of burnout, when high levels of exposure to moral injury occurred (ibid). Martin et al^20^ found perfectionism to significantly predict emotional exhaustion (β = 0.55, p = <.001) and depersonalisation (β = 0.18, p = .006) in physicians, while Biggs, McKay and Shanmugam^21^ showed SPP to be associated with burnout (β = 0.36 p = .001) in physiotherapy students, suggesting perfectionism renders the person vulnerable to burnout. Against this backdrop, we argue the tendency to achieve perfection in the workplace adversely affects performance, placing individuals at risk of burnout.

The intersection between perfectionism and moral injury is of particular interest, especially in how SPP might interact with moral injury. One plausible hypothesis is SPP, the dimension of perfectionism aimed at meeting the standards set by significant others, could lead to feelings of betrayal, subsequently triggering moral injury. To the authors’ knowledge, perfectionism and moral injury have not been investigated together. However, it seems logical SPP could serve as a predictor of moral injury. The relationship between perfectionism, burnout and moral injury has also not been explored. Given the established associations of both perfectionism and moral injury with burnout in the literature^13,20^, and considering perfectionism predicts moral injury, it is reasonable to infer a connection between these variables. These three variables, despite limited research, each demonstrated statistical significance in healthcare professionals worldwide, with no data, however, pertaining to perfectionism, moral injury and burnout in UK-based physiotherapists. This study aims to be the first to comprehensively examine these three variables among UK-based physiotherapists. We anticipate, firstly, a significant association between perfectionism and moral injury with burnout. Secondly, perfectionism, we expect, will predict moral injury, and in turn burnout. Therefore, this investigation aims to understand the nature and extent of the relationship between perfectionism, moral injury and burnout in UK-based Physiotherapists.

## Method

### Methodology

In this research, we followed the “Strengthening the Reporting of Observational Studies in Epidemiology” (STROBE) guidelines (von Elm et al. 2008). Our study specifically utilized the cross-sectional study version of the STROBE Checklist, incorporating 22 key points for survey design and result reporting, as detailed in Supplementary Table 1.

### Measures

Participants completed a Microsoft (MS) Forms survey which consisted of 12 questions related to demographics and work environment as well as three validated scales, measuring perfectionism, moral injury, and burnout in the profession. Perfectionism was measured via the Multidimensional Perfectionism Scale Short Form^4^ (MPS-SF), which measures the aforementioned dimensions of SOP and SPP. Moral Injury was measured via Moral Injury Symptoms Scale-Healthcare Professionals Version (MISS-HF). ^22^ Due to limitations on the website hosting the survey, the Likert scale used to measure moral injury required modification from 10 levels to 7. Finally, Burnout was measured via the Shirom-Melamed Burnout Questionnaire^23^ using the domains of EE, PF, and CW. The Likert scale used to measure burnout also required modification to fit the website hosting the survey from a Likert of 7 to 5.

### Procedure

Participants found the survey through links on posts from the investigators’ social media accounts or through recruitment emails. By clicking on the link, participants are directed to the MS Forms website where they can complete the questions.

### Ethics

Informed consent was implied in the participation of the survey. The survey preamble explicitly stated the purpose of the study, assured anonymity and confidentiality, and emphasized the voluntary nature of participation. Further information was provided in a participant information sheet. In the first question, participants were informed that by proceeding with the survey, they were providing consent to utilize their responses for research purposes. If they opted yes, they go forward with the completion of the survey and if they opted no, they would exit the survey. The start date of recruitment was 25 November 2022 and the end date 30 April 2023. They were informed of contact details for the research team and potential emotional impact. Post-survey, resources for mental health support were provided. Ethical approval was obtained from Glasgow Caledonian University’s School of Health and Life Sciences (HLS/PSWAHS/22/017).

### Data Analysis and Study Size

Preliminary, descriptive, and correlation analysis were conducted in SPSS 26.0. Scores were categorised as low, medium, and high by dividing the highest possible score for each measure into three parts. Preliminary analysis comprised out-of-range values, univariate, and multivariate normality, and reliability. Two-step structural equation modelling (SEM) was undertaken in AMOS 26.0. ^24^ Maximum likelihood estimation was employed to access the goodness-of-fit and parameters of the statistical model. The model included four interrelated latent variables: SSP, SOP, moral injury, and burnout. Burnout was indicated by three dimensions (EE, PF, and CW) and moral injury, SOP and SPP were indicated by their respective items. This approach enabled the power analysis calculation of the model that showed 95% confidence interval with a 5% margin of error within the sample and shows representation of the UK Physiotherapist population. The Structure Equation Model assessment progressed sequentially, starting with measurement model evaluation, followed by confirmatory factor analyses. Subsequently the theorised structural relationships were evaluated. Conventional criteria of approximate markers of acceptance (χ2/df ratio < 3.00, IFI and CFI > 0.90, RMSEA < 0.08) and excellent fit (χ2/df ratio < 2.00, IFI and CFI > 0.95, RMSEA < 0.06) as recommended by Marsh et al.^25^

## Results

### Participants

Between October 2022 and February 2023, 402 UK-based physiotherapists completed an online survey. Participants were categorised as: Private Physiotherapists (9%), NHS Public Physiotherapists (87%), Sports (<1%), Academia (3%) and Other (<1%) (see Table 1). Participants worked in a range of clinical and non-clinical physiotherapy environments and specialties, including inpatient, outpatient, and higher education. Participants reported different duration of employment, with 23% working 2-5 years, 21% over 10 years, and 19% 12-24 months in their current position. See table 2 for an overview of demographics, including gender, ethnicity, and education. Recruitment occurred through convenience sampling, advertising through social media and through emails with an invitation to complete the survey. Inclusion criteria included participants 21 years of age or over and work as a physiotherapist in the United Kingdom.

**Table 1:** Categorisation of Physiotherapy Positions.

| Categorisation of Participants | Number within the field | Percentage |
| --- | --- | --- |
| Band 5 NHS | 52 | 13 |
| Band 6 NHS | 91 | 23 |
| Band 7 NHS | 129 | 32 |
| Band 8a | 55 | 14 |
| Band 8b | 17 | 4 |
| Band 8c | 3 | <1 |
| Band 8d | 1 | <1 |
| Private Practice | 35 | 9 |
| Sports-Based | 3 | <1 |
| Lecturer | 5 | 1 |
| Senior Lecturer | 7 | 2 |
| Reader | 1 | <1 |
| Other | 3 | <1 |

**Table 2.** Frequency and Percentage of the Key Demographics.

| Demographics |  |
| --- | --- |
| **Gender** |  |
| Female | 330 (82%) |
| Male | 67 (17%) |
| Non-binary | 1 (<1%) |
| Prefer not to say | 4 (1%) |
| **Age** |  |
| 21-24 years | 29 (7%) |
| 25-29 years | 40 (10%) |
| 30-34 years | 52 (13%) |
| 35-39 years | 71 (18%) |
| 40-44 years | 77 (19%) |
| 45-49 years | 55 (14%) |
| 50+ years | 78 (19%) |
| **Ethnicity** |  |
| Asian or Asian British | 28 (7%) |
| Black, Black British, Caribbean or African | 8 (2%) |
| Mixed or Multiple Ethnic Groups | 18 (4%) |
| White | 348 (87%) |
| **Education** |  |
| BSc | 191 (48%) |
| MSc | 164 (41%) |
| PhD | 22 (5%) |
| Professional Doctorate | 1 (<1%) |
| DPT | 8 (2%) |
| PgDip | 13 (3%) |
| MRes | 3 (1%) |

Bachelor of Science. MSc = Master of Science. PhD = Doctor of Philosophy. DPT = Doctor of Physiotherapy. PgDip = Postgraduate Diploma. MRes = Master of Research.

### Preliminary Analyses, Descriptive Statistics, and Bivariate Correlations

Table 3 presents the average (mean), variance (standard deviation), correlation (Pearsons), and level of significance for each total of the MPS-SF (excluding other oriented perfectionism), MISS-HF, and SMBQ domains. For the SOP and SPP domains of the MPS-SF, the highest total score possible is 35. The highest total score possible for the MISS-HF is 70. SMBQ highest possible total score is 70 while the highest scores for each domain include 42 for PE, 21 for EE, and 35 for CW. All values lay within the expected range of the scale. No data was missing due to the requirement in the survey for participants to complete each question. Data for all scales fit normal distribution models of absolute skewness and absolute kurtosis.

**Table 3.** Mean, standard deviation, and correlations of the three outcome measures.

|  | 1.SOP | 2.SPP | 3.Moral Injury | 4.Physical Exhaustion | 5.Emotional Exhaustion | 6.Cognitive Weariness | 7.Total Burnout | Mean | SD |
| --- | --- | --- | --- | --- | --- | --- | --- | --- | --- |
| 1 |  | .576** | .291** | .535** | .124* | .240* | .108* | 25.2 | 6.8 |
| 2 |  |  | .482** | .540** | .346** | .397** | .336** | 22.4 | 6.7 |
| 3 |  |  |  | .504** | .461** | .432** | .559** | 34.7 | 8.4 |
| 4 |  |  |  |  | .473** | .641** | .886** | 21.5 | 5.8 |
| 5 |  |  |  |  |  | .514** | .727** | 7.3 | 7.3 |
| 6 |  |  |  |  |  |  | .873** | 15.2 | 5 |
| 7 |  |  |  |  |  |  |  | 44.1 | 12 |

*Note.* Cronbach Alpha - SOP .84, SPP .90, Moral Injury .74, Burnout .94 Sig level \* .05. \*\* .01. SOP= socially oriented perfectionism. SPP = socially prescribed perfectionism.

All correlations between the total and domain total scores for each scale were found to be significant (see table 3). Table 4 highlights the percentages of the sample by category level in outcome scores. As can be seen, 95% of physiotherapists experienced moderate-to-high levels of SOP, 92% had moderate-to-high levels of SPP, 90% had moderate-to-high levels of moral injury, and 96% had moderate-to-high levels of burnout.

**Table 4.**
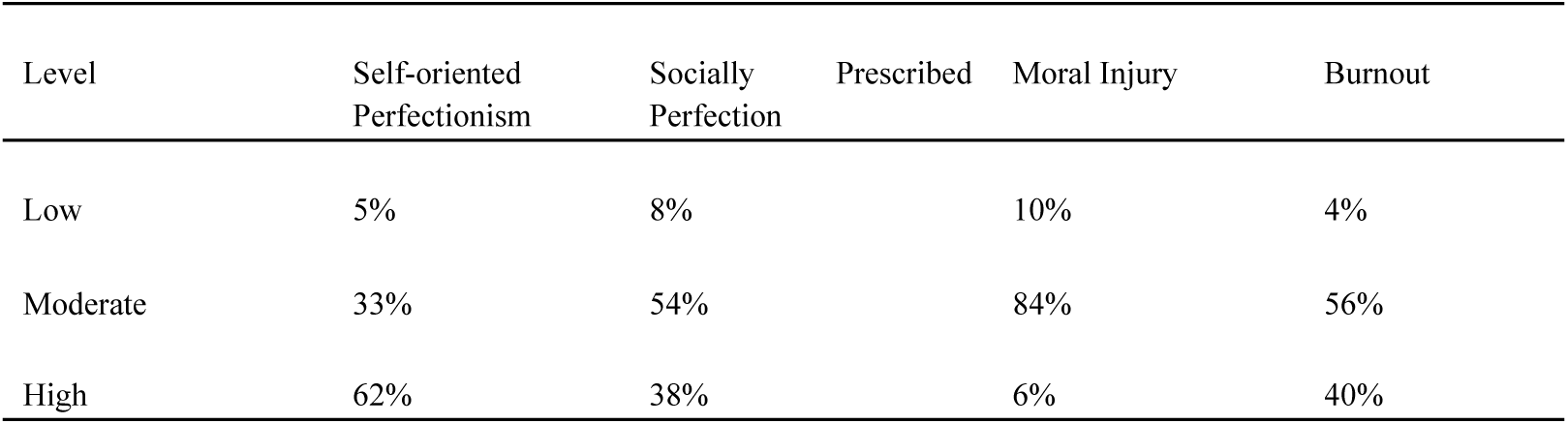
Population Percentage by Category Level Proportion in Each Outcome Measure.

### Structural Equation Modelling

Confirmatory factor analyses indicated acceptable-to-excellent fit for the measurement model (χ2/df ratio = 2.25, IFI = 0.92, CFI = 0.92, RMSEA = 0.06 CI 0.06-0.07). Composite reliabilities (ρc) supported the measurement model: SOP = 0.90; SPP = 0.84; Moral Injury = 0.74 and Burnout = 0.94. Structural Equation Modelling also indicated an acceptable-to-excellent fit (χ2/df ratio = 2.25, IFI = 0.92, CFI = 0.92, RMSEA = 0.06 CI 90% 0.06-0.07, SRMR = .067). All residual covariances were < 2.0 Overall, the model explained 62% variance in burnout. Parameters are displayed in Figure 1.

**Figure 1.**
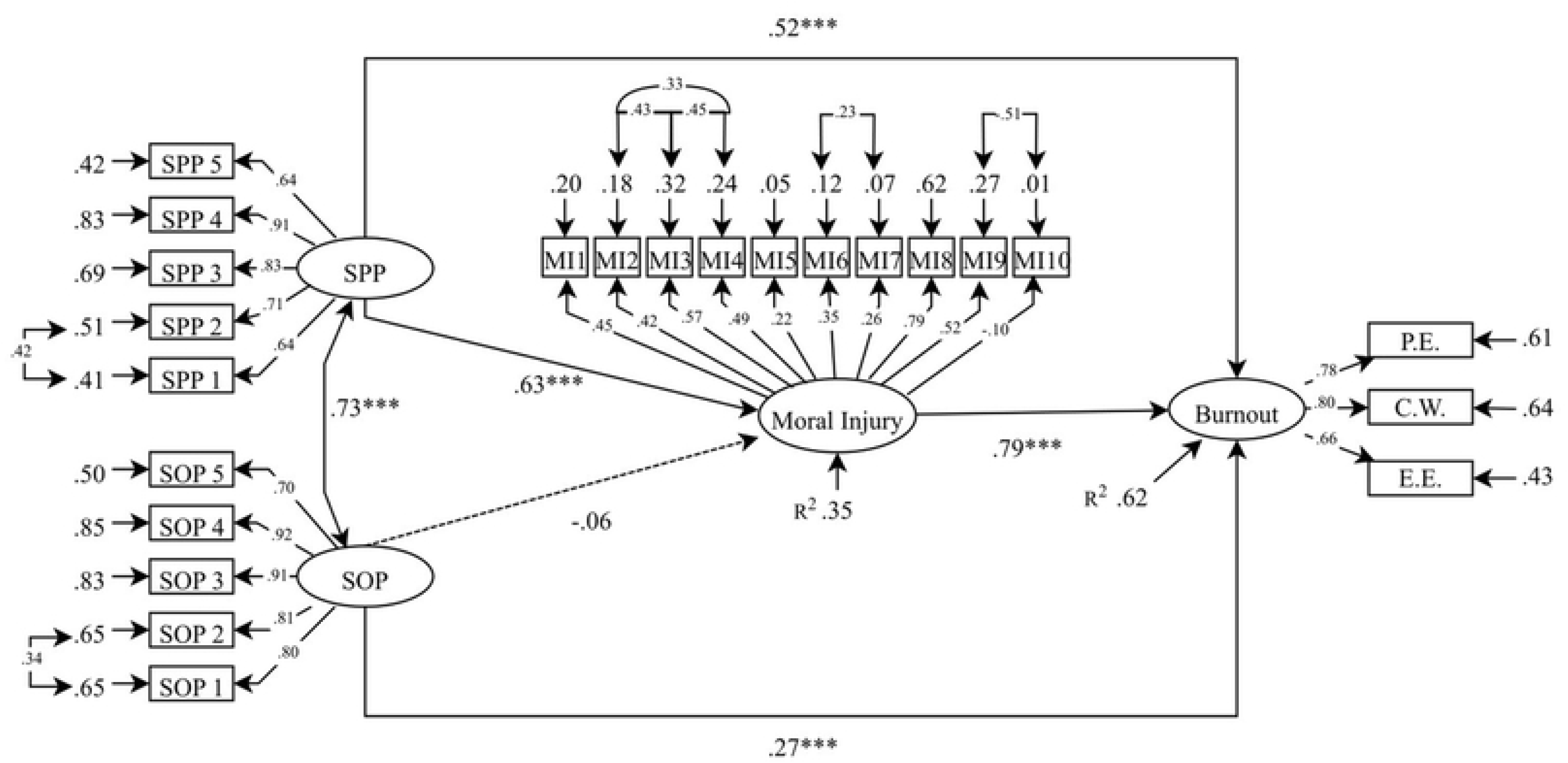
Structural equation model to show the relationship between perfectionism, moral injury and burnout.

## Discussion

Our study sought to explore the relationship between perfectionism, moral injury, and burnout in physiotherapists based in the UK, through both correlation analyses and SEM techniques. We found a substantial proportion of variability in burnout can be accounted for by perfectionism and moral injury. As expected, we uncovered noteworthy positive relationships between SPP-Burnout, SPP-Moral Injury, and Moral Injury-Burnout. Notably, a statistically significant pathway emerged from perfectionism (SPP) to moral injury, and subsequently to burnout. This underscores two crucial findings. Firstly, perfectionism and moral injury exhibited meaningful associations with burnout. Secondly, perfectionism served as a precursor to moral injury, which in turn predicted burnout.

In essence, our study not only establishes the existence of significant relationships among perfectionism, moral injury, and burnout but also shows a sequential pattern wherein perfectionism sets the stage for moral injury, ultimately contributing to the manifestation of burnout.

### Burnout in Physiotherapy

96% of physiotherapists in our sample suffered from moderate-to-high levels of burnout, presenting a pervasive challenge. Implications of these scores are concerning for several reasons. When considering our study and the research by Donohoe et al^18^ who found 40% of physiotherapists suffered moderate to high burnout levels, our burnout levels reveal a marked increase. These heightened levels present immediate challenges, with potential depletion of the physiotherapy workforce intensifying pressure on existing staff. Secondly, burnout has been linked to higher turnover rates among staff.^26^ Given the existing shortage of physiotherapists in the UK, the high levels of burnout in this study may be a contributing factor to a significant exodus from the profession. ^27^ Our findings not only underscore the severity of burnout-related issues but also offer a potential explanation for the scarcity of physiotherapists in the UK.

Individuals can encounter challenges, such as impaired executive function, compromised episodic and working memory, and diminished task attention when facing burnout.^28^ With appropriate intervention, these issues can be reversed.^29^ Our examination of burnout dimensions reveals both physical and emotional exhaustion scores on average fall between medium and high. In a study by Brand et al^30^, 21.1% of health professionals with high burnout scores experienced insomnia. Given sleep deprivation adversely affects attention, working memory, long-term memory, and decision-making^31^, it may explain why these symptoms have been reported as risk factors in burnout research.^29^ It is then logical to infer insomnia might be prevalent among UK physiotherapists, given the burnout scores observed in our sample. As a result, despite experiencing exhaustion, achieving restful sleep may be elusive due to the coexistence of insomnia. If physiotherapists are indeed dealing with insomnia, the quality of care provided may be compromised, posing additional health risks to their patients.

Given that this study represents one of the first to capture burnout in a sample of UK physiotherapists, with such stark results, urgent and comprehensive efforts are required to mitigate the impending risks to staff, patients, and the broader society. Understanding the reasons behind the increase in burnout rates requires further exploration.

### Perfectionism and Burnout

On average, we observed moderate to high levels of SOP and SPP in our physiotherapy population. Correlational analysis reveals a connection between SPP, SOP, and moral injury and burnout. Further analysis indicates a positive association between both SPP and moral injury with burnout. The self-induced external perfectionistic standards appear to be the closest predictor of burnout. In practical terms, this connection could manifest when a physiotherapist achieves a set goal, but external hierarchical peers may fail to provide sufficient feedback that meets the perfectionistic standards of the feedback-reciprocal physiotherapist. Consequently, the physiotherapist, in such cases, becomes vulnerable to experiencing elevated levels of burnout.

Our findings align with previous research identifying SPP as a stronger predictor of burnout compared to SOP.^21^ Recently, Biggs McKay and Shanmugam^21^ found a significant association between SPP and burnout in UK physiotherapy students, suggesting a consistent trend across different professional stages. Our findings also contribute to a growing body of evidence linking perfectionism, particularly SPP, to adverse health implications. SPP demonstrates a stronger direct relationship with clinically diagnosed conditions, including affective disorders, eating disorders, and narcissistic personality disorder.^5^ The relationship between SPP and burnout observed in our sample underscores the importance of recognising and addressing maladaptive perfectionism tendencies, as they can have significant implications for individual health and well-being.

### Moral Injury and Burnout

Mantri et al^32^, in their sample of 1831 health professionals, found a significant relationship between moral injury and burnout. The authors understood the surge in rates of moral injury found in their data to be in response to the pandemic. On average, our sample consisted of moderate levels of moral injury. Further analysis revealed a significant correlation between burnout and moral injury, similar to previous research^13^ and the understanding that burnout might not be due to pandemic related factors alone.^32^ SEM helped us establish a significant association, with moral injury predicting burnout (β 0.78). Our study stands as the first to uncover such an association in the physiotherapy profession. Building on the work of Mantri et al^32^, we found moral injury can predict burnout, and when combined with perfectionism, it explains 62% of the variance in burnout scores.

Our study is the first to establish a relationship between SPP and moral injury (β 0.54). We theorise the external standards set by significant others could trigger a sense of betrayal, subsequently leading to moral injury. Our results support this association. Considering the mean SPP score in our sample is moderate to high, it implies that physiotherapists are striving for standards of perfection as influenced by their peers. However, this pursuit may render them susceptible to moral injury if perceived betrayal from significant peers, such as line managers or superiors, occurs. This vulnerability likely predisposes physiotherapists to a risk of elevated levels of burnout.

### Perfectionism, Moral Injury and Burnout’s Pathway

This study marks the first instance where the relationship between perfectionism and burnout is elucidated from the perspective of a betrayal of values. The SEM results extend the findings of Martin et al^20^ and Testoni et al^13^, underscoring the susceptibility to burnout exhibited by SPP physiotherapists when confronted with moral injury. SPP was not the only domain on the MPS-SF to have a relationship with burnout. A relationship between SOP and burnout, observed through SEM, was not significantly associated by moral injury, despite a modest association between the two (β 0.27). Our research suggests SOP has a detrimental relationship with burnout, explaining some of the variance in scores. Logically, it becomes evident that SOP correlates with the three dimensions of burnout (see Table 3), exposing individuals to heightened exhaustion and cognitive weariness as they strive to meet the impeccable standards they set for themselves.

### Practical Implications

Our research underscores the importance of the interplay between perfectionism and moral injury for physiotherapists and their line managers. Although our research focuses on a physiotherapy population, high burnout levels are likely not unique to the profession and it can be expected to be found across other health professionals. Burnout, as such, can be considered a ‘wicked’ problem and of great concern to the sustainability of the NHS workforce. Challenges such as workload pressures, budget constraints, staff shortages and patient wait times would be and are increasing due to the sustainability of the practitioners in physiotherapy. Strategies need to be developed to manage risk factors to burnout and support those impacted by the syndrome at a national level. Creating these strategies will likely benefit from a collaborative effort between key stakeholders interested in supporting the mental health and job satisfaction of health care workers, including the healthcare workers affected. For example, the use of co-creation methodologies may facilitate interventions that are realistic, person-centred, and achievable.

### Limitations and Future Research Directions

Although our study can explain around 60% of the variance of burnout in physiotherapists, other influences of the remaining 40% remain unknown. We also cannot fully understand the experience of burnout for physiotherapists through their survey response. For example, some physiotherapists may leave the workforce due to burnout while some may persevere or use unknown coping strategies to continue in their post. Understanding the experience of burnout would help inform the creation of strategies. It is also unknown if the adaptation of the Likert scales used in the survey reduced the level of detail in participant scores. For example, with fewer points on the scale, we may have missed small differences in burnout level between participants. However, it is also unclear how much this other detail would have contributed to our understanding and evaluation of the problem.

Naturally with most surveys, participant answers are subjective, vulnerable to over or underestimation, and only offer us a time snapshot of perfectionism, moral injury, and burnout in UK-based physiotherapists. We cannot understand if these rates are static, vary over the course of the year, or if they still remain high at nearly one year after the start of data collection. Future research may wish to explore how burnout changes over time, with intervention, and across different health professions. Nonetheless, the results may be generalisable to an extent as burnout can be observed in physicians and medical doctors and thus the path of perfectionism-moral injury-burnout may also be present in these populations too.^20^

In this publication, we focused on the role of SOP and SPP and their relationship with moral injury and burnout. It may also be of interest to understand the role of OOP. OOP describes behaviours related to the placement of high standards on others. As we know these standards negatively impact physiotherapists with expectations of perfection placed upon them by others, an understanding of the prevalence of OOP may help inform strategies to reduce toxic behaviours in team dynamics and the wider profession. It may also prove informative to explore sleep hygiene practices and sleep quality in the profession to better understand the connection of insomnia with burnout.

## Conclusion

This study offers valuable insights into the intricate dynamics among perfectionism, moral injury, and burnout, emphasising that moral injury acts as a pivotal mediator in the relationship between SPP and burnout. The collective influence of perfectionism and moral injury emerges as a robust predictor of burnout in physiotherapists. Importantly, our findings contribute to the growing body of evidence highlighting the health risks associated with SPP across diverse populations.

The unprecedented levels of burnout observed in UK-based physiotherapists, as revealed by our study, underscore a critical concern with far-reaching implications. This high prevalence of burnout has the potential to impact not only the well-being of physiotherapy professionals but also poses risks to patient care, staff dynamics, and the broader society. The implications extend beyond individual practitioners to the healthcare ecosystem as a whole.

In essence, this study highlights the urgent need for targeted and systemic interventions along with comprehensive strategies to address the pervasive challenges posed by perfectionism, moral injury, and burnout within the field of physiotherapy. Acknowledging and actively mitigating these issues is imperative for fostering a resilient and sustainable healthcare workforce, ultimately enhancing the quality of patient care and safeguarding the well-being of both physiotherapists and the communities they serve.

## Data Availability

Data cannot be shared publicly because of confidentiality and anonymity reasons and complying with the remit of ethical approval. Data are available from the Glasgow Caledonian University Institutional Data Access / Ethics Committee (contact via the primary author – Dr Sivaramkumar Shanmugam) for researchers who meet the criteria for access to confidential data.

## Acknowledgements

This work praises the time and effort spent by the Physiotherapists in the UK. The project did not acquire funding to undertake.

## Reference List

(1) Flett GL, Hewitt PL., Perfectionism: Theory, research, and treatment. American Psychological Association: 2002. Chapter 1, Perfectionism and maladjustment: An overview of theoretical, definitional, and treatment issues; p. 5–31.

(2) Frost RO, Marten P, Lahart C, Rosenblate R. The dimensions of perfectionism. Cognitive therapy and research 1990;14:449–468.

(3) Hewitt PL, Flett GL. Dimensions of perfectionism in unipolar depression. J Abnorm Psychol 1991;100(1):98

(4) Cox BJ, Enns MW, Clara IP. The multidimensional structure of perfectionism in clinically distressed and college student samples. Psychol Assess 2002;14(3):365.

(5) Flett GL, Hewitt PL, Nepon T, Sherry SB, Smith M. The destructiveness and public health significance of socially prescribed perfectionism: A review, analysis, and conceptual extension. Clin Psychol Rev 2022;93:102130.

(6) Chartered Society of Physiotherapy. 2017. CSP facts and figures. Available: https://www.csp.org.uk/about-csp/who-we-are/facts-figures [Accessed 09.01/2024].

(7) Shay J. Achilles: Paragon, Flawed Character, or Tragic Soldier Figure? The Classical Bulletin 1995;71(2):117

(8) Shay J. Moral injury. Psychoanalytic psychology 2014;31(2):182.

(9) Brock RN, Lettini G. Soul repair: Recovering from moral injury after war.: Beacon Press; 2012.

(10) Dean W, Talbot S, Dean A. Reframing clinician distress: moral injury not burnout. Federal Practitioner 2019;36(9):400.

(11) Whitehead PB, Herbertson RK, Hamric AB, Epstein EG, Fisher JM. Moral distress among healthcare professionals: Report of an institution-wide survey. Journal of nursing scholarship 2015;47(2):117–125.

(12) Čartolovni A, Stolt M, Scott PA, Suhonen R. Moral injury in healthcare professionals: A scoping review and discussion. Nurs Ethics 2021;28(5):590–602.

(13) Testoni I, Brondolo E, Ronconi L, Petrini F, Navalesi P, Antonellini M, et al. Burnout following moral injury and dehumanization: A study of distress among Italian medical staff during the first COVID-19 pandemic period. *Psychological Trauma: Theory, Research*, Practice, and Policy 2022; 15(Suppl 2).

(14) Williams ES, Rathert C, Buttigieg SC. The personal and professional consequences of physician burnout: a systematic review of the literature. Medical Care Research and Review 2020;77(5):371–386.

(15) Maslach, C., Jackson, S. (1984), Burnout in organization settings. Applied Social Psychology Annual, 5(1), 133–153.

(16) Lundgren-Nilsson Å, Jonsdottir IH, Pallant J, Ahlborg G. Internal construct validity of the Shirom-Melamed burnout questionnaire (SMBQ). BMC Public Health 2012;12:1–8.

(17) Gustafsson H, Kenttä G, Hassmén P. Athlete burnout: An integrated model and future research directions. International Review of Sport and Exercise Psychology 2011;4(1):3–24.

(18) Donohoe E, Nawawi A, Wilker L, Schindler T, Jette DU. Factors associated with burnout of physical therapists in Massachusetts rehabilitation hospitals. Phys Ther 1993;73(11):750–756.

(19) Bejer A, Domka-Jopek E, Probachta M, Lenart-Domka E, Wojnar J. Burnout syndrome in physiotherapists working in the Podkarpackie province in Poland. Work 2019;64(4):809–815.

(20) Martin SR, Fortier MA, Heyming TW, Ahn K, Nichols W, Golden C, et al. Perfectionism as a predictor of physician burnout. BMC Health Services Research 2022;22(1):1425.

(21) Biggs, P. D, McKay, J., Shanmugam, S. Academic Burnout and Perfectionism in UK-based Physiotherapy Students. Advances in Health Sciences Education 2024 Forthcoming.

(22) Mantri S, Song YK, Lawson JM, Berger EJ, Koenig HG. Moral injury and burnout in health care professionals during the COVID-19 pandemic. J Nerv Ment Dis 2021;209(10):720–726.

(23) Shirom A, Melamed S. A comparison of the construct validity of two burnout measures in two groups of professionals. International journal of stress management 2006;13(2):176.

(24) Anderson JC, Gerbing DW. Structural equation modeling in practice: A review and recommended two-step approach. Psychol Bull 1988;103(3):411.

(25) Marsh HW, Hau K, Wen Z. In search of golden rules: Comment on hypothesis-testing approaches to setting cutoff values for fit indexes and dangers in overgeneralizing Hu and Bentler’s (1999) findings. Structural equation modeling 2004;11(3):320–341.

(26) Willard-Grace R, Knox M, Huang B, Hammer H, Kivlahan C, Grumbach K. Burnout and health care workforce turnover. The Annals of Family Medicine 2019;17(1):36–41.

(27) Dolton, P., Nguyen, D., Castellanos, M., & Rolfe, H. 2018. BREXIT and the Health & Social Care Workforce in the UK. Available: https://www.niesr.ac.uk/wp-content/uploads/2021/10/Report-Brexit-Health-and-Social-Care-Workforce-Full-Report-4.pdf [Accessed 07/01/2024]

(28) Jonsdottir IH, Nordlund A, Ellbin S, Ljung T, Glise K, Währborg P, et al. Working memory and attention are still impaired after three years in patients with stress-related exhaustion. Scand J Psychol 2017;58(6):504–509.

(29) Bayes A, Tavella G, Parker G. The biology of burnout: Causes and consequences. The World Journal of Biological Psychiatry 2021;22(9):686–698.

(30) Brand S, Beck J, Hatzinger M, Harbaugh A, Ruch W, Holsboer-Trachsler E. Associations between satisfaction with life, burnout-related emotional and physical exhaustion, and sleep complaints. The World Journal of Biological Psychiatry 2010;11(5):744–754.

(31) Alhola P, Polo-Kantola P. Sleep deprivation: Impact on cognitive performance. Neuropsychiatric disease and treatment 2007;3(5):553–567.

(32) Mantri S, Song YK, Lawson JM, Berger EJ, Koenig HG. Moral injury and burnout in health care professionals during the COVID-19 pandemic. J Nerv Ment Dis 2021;209(10):720–726.

